# Surgical diagnoses and post-operative outcomes of intestinal obstruction among adults at regional referral hospitals in Dar-es-Salaam, Tanzania: a prospective, observational hospital-based study

**DOI:** 10.1101/2023.12.31.23300678

**Authors:** Shadrack Samwel Mponzi, Wambura Boniphace Wandwi, Naboth Almasi Mbembati, Kelvin Melkizedeck Leshabari

## Abstract

**Background:** There is limited available published information regarding surgical diagnoses and post-operative outcomes of intestinal obstruction among adult patients in East African hospitals. The observation is despite available anecdotal evidence of the condition to be among the commonest surgical emergencies in the area.

**Objective:** To assess the surgical diagnoses and outcomes of intestinal obstruction among adult patients treated at the regional referral hospitals in Dar-es-salaam - Tanzania.

**Methods & Findings:** This was a prospective, observational, hospital-based study. Data were collected using a pre-validated Clinical Research Form (CRF). All adult patients with post-operative surgical diagnoses of intestinal obstruction at Amana, Mwananyamala and Temeke hospitals in Dar es Salaam were the target population. Data were analyzed using a generalized linear model via SAS version 9.7. Multivariable logistic regression model was the final fitted model. Intra-operative findings (surgical diagnoses) of intestinal obstruction was an outcome variable. Unless otherwise stated, an α-level of 5% was used as a limit of type 1 error in findings. The study analysed an average of 1411 patients-days of follow-up. Participants’ median age and duration of hospital stay were 47 (IQR: 35-67) years and 4 (IQR: 3-6) days respectively. Intra-operative findings included adhesions (aOR=5.66), abdominal tumors (aOR=1.028), hernia (aOR=2.04) and volvulus (aOR=4.2). Moreover, 12 (5.26%) clients died and 20 (9%) had surgical sites infection. No statistically significant difference of hospital on surgical outcomes (χ^2^ test value = 4.992; df = 10).

**Conclusion:** Adhesions was the commonest intraoperative cause of intestinal obstruction in this study population. One-in-twenty of all followed-up clients died. Significant proportion of patients had evidence of post-operative complications.

## INTRODUCTION

Intestinal obstruction can be described as either complete or partial occlusion, of intestinal contents through the alimentary canal, from the level of the small bowel and beyond, brought about by either mechanical (dynamic) or functional (adynamic) factors [1]. Complete intestinal obstruction refers to a situation, where neither digested food particles, nor gas (flatus), can pass through the bowel, distal to the site of obstruction [1]. Partial intestinal obstruction occurs when gas and/or scanty fecal matter can pass through to distal destination(s) from the site of obstruction [1]. Globally, intestinal obstruction is a surgical emergency with high fatality rates (up to 78%) when unattended [2-4]. There is paucity of findings from developing countries, especially Tanzania; that address the burden, clinical characteristics, causative factors as well as outcomes of bowel obstruction [3, 5-8]. The few available ones, are mainly either targeting specific pathology associated with intestinal obstruction [7], or biased by the mechanism of obstruction [8]. Thus, we decided to fill the current gap, by *assessing surgical diagnoses and post-operative outcomes, of intestinal obstruction, irrespective of the root cause or mechanism producing the obstruction process, among adults residing in a typical metropolitan setting of Africa*.

Several clinical characteristics are associated with intestinal obstruction in literature [9-11]. On symptomatology, they include abdominal pain, distension, and absence of passage of flatus/feces, vomiting and nausea [9, 10]. Likewise, abdominal tenderness, abdominal distension and increased bowel sounds have been reported as frequent signs in literature [11]. Abdominal wall hernias rank first among aetiological factors responsible for intestinal obstruction reported from developing countries while intra-abdominal adhesions are the commonest insults reported in developed countries [3, 11-13]. Additionally, intra-abdominal wall hernias and intra-abdominal adhesions are the commonest etiologies of mechanical bowel obstruction in young and adults while obstructive tumors and torsion of the bowel are the commonest etiological factors responsible for mechanical bowel obstruction in the elderly [1]. However, most of the studied clinical features for intestinal obstruction found in literature referred to mechanical intestinal obstruction; with little references to functional intestinal obstruction [3, 11, 14, 15]. In the current study, clinical characteristics were determined with reference to the mechanism(s) of obstruction.

Management of bowel obstruction depends on accurate and timely diagnosis and treatment plans. Dichotomized model of bowel obstructions into mechanical (dynamic) and functional (adynamic) intestinal obstruction; assists in a number of clinical activities ranging from etiological classification to diagnostic as well as therapeutic options. Specifically, mechanical bowel obstruction clinically refers to physical luminal occlusion that takes place either within the lumen (worms, gallstones, fecal matter, foreign bodies), in the intestinal wall (tumors, strictures) or outside the intestinal wall (adhesions/bands, hernias and volvulus) [16]. Functional (adynamic) bowel obstruction reflects systemic impairment that results in loss of peristaltic or non-propulsive movement if any, for propelling the food matter distally which can be either due to defective intestinal smooth muscles (myogenic) or defective nerve supplying the muscles (neurogenic) on clinical grounds [16]. Occasionally, the model is deficient in cases of mixed etiologies. For instance, outlet obstruction (anorectal) prevents defecation and the causes also range from mechanical to functional bowel obstruction. Besides, for temporal association between an insult and a surgical diagnosis to be realized, clinicians ought to prospectively assess patients, least of that the findings thereof are likely to be accompanied by different forms of biases. Thus, we designed a prospective observational hospital-based study, to assess the diagnostic as well as outcomes of surgical interventions among adult patients, with symptoms and signs suggestive of intestinal obstruction.

## METHODS

### Study Design and settings

We designed a prospective, hospital-based follow-up study in Dar es Salaam public regional referral hospitals. Specifically, the study took place at Amana, Mwananyamala and Temeke regional referral hospitals. Recruitment started at the operating theatre prior to surgical opening. Follow-up was up to (and including) time of discharge from the ward post-operatively. The decision to conduct the study at the specified sites included the fact that they included a rich mix of all demographic sects of the population residing in Dar es Salaam. Dar es Salaam is the business capital of Tanzania. It is situated on the East African coastal shores. The demographic subsets of Dar es Salaam encompasses diverse African population groups ranging from Bantus to Afro-Arabic and Afro-Indo-Persian people. Otherwise, Dar es Salaam is among the fastest rapidly growing cities, not only in Africa but worldwide. According to UN department of Economic and Social Affairs, although Dar es Salaam is placed at no.11 out of 15 rapidly growing cities in the world by 2035 [17], by sheer proportional increase by percentage, it is in fact the top most of all the cities in terms of the gross number increase of population statistics in the same predicted time frame [17]. By far, Dar es Salaam is estimated to be the 2^nd^ largest city in population size on earth, only behind Lagos, Nigeria come the year 2100 [18]. However, Lagos metropolitan is mainly inhabited by population currently ethnographically characterized as Bantus, leaving Dar es Salaam as the true megacity with likelihood to represent nearly all diverse demographic subsets of Africa.

### Study Population and Target Population

Adult patients with surgical diagnosis of intestinal obstruction were eligible to participate into this study. Specifically, we targeted adult patients (aged ≥ 18 years), who underwent surgery due to presenting symptoms and signs suggestive of *acute intestinal obstruction* at Dar-es-salaam regional referral hospitals.

### Inclusion and Exclusion criteria

We included patients who were at least 18 years old. Besides, for all included patients, a provisional surgical diagnosis of acute intestinal obstruction had to be made either clinically or radiologically at the study settings. Conversely, patients referred from other centers to the study sites with proven failed surgery (laparotomy) earlier were excluded from our current study follow-up and analysis.

### Data collection procedures

Data collection started at surgical theatre. All adult patients who were scheduled and later underwent laparotomy due to symptoms and signs suggestive of *acute intestinal obstruction* were invited to participate, had to sign their written informed consent (after a brief description about the study goal, nature, risks vs. benefits of participation as well as to whom should the correspondence – when needed, be addressed), and followed-up at Amana, Mwananyamala & Temeke regional referral hospitals. Initial clinical data (mainly demographics – age, gender; comorbidity stata – past and present medical/surgical history, including their current drugs history); as well as intra-operative findings (i.e. surgical diagnosis) were recorded in a pre-designed *clinical report form*. Post-operative interventions in the ward, included wound status, length of hospital stay and mortality for each patient were also followed-up and subsequently analysed. Post-operative morbidity/mortality by type was also recorded during hospital stay post-operatively. All documented details during data collection, data processing and data analysis were carefully managed by the principal investigator.

### Data management and Analysis

Information once collected on the pre-validated clinical sheet was entered on computer database. Data was then stored in PI’s computer. Initial data exploration was done, and mainly involved data summarization, trend in data as well as consistency in data structure. Quantitative data were all skewed and hence had to be summarized using median (with corresponding inter-quartile range) while qualitative data were summarized using frequency and proportions. Data analysis was done via SAS software version 9.7. Main data analysis involved fitting of a generalised linear model after necessary model assumptions validation. Specifically, we fitted a multivariable binary logistic regression model as the main statistical tool to answer our research hypothesis. Model validation for the fitted linear model considered both tests for *homoscedasticity, autocorrelation, multicollinearity, normality* as well as *linearity* assumptions. During univariate analyses, an α-level of 0.2 was set as a limiting value for significance. Besides, for each variable that was included in the multivariable analysis, a significance level of 5% was used as a limit of type I error. The null hypothesis in this study was “*there is no difference in outcomes between patients treated for intestinal obstruction at Dar es Salaam regional referral hospitals*”.

## RESULTS

The study analysed an average of 1411 patients-days of follow-up from Amana, Mwananyamala and Temeke regional referral hospitals covering the period from December 2021 to July 2022. Specifically, they included 51 (22%) patients from Amana, 95 (42%) from Mwananyamala and 82 (36%) from Temeke hospitals. Their baseline characteristics included male: female = 3:1 (males: n=172, 75.4%). Median age of study participants was 34 (IQR: 27-45) years. Likewise, the median duration of hospital stay was 4 (IQR: 3-6) days. During the study period, 12 clients died, making the crude mortality rate of 5.3%. Details about frequency and contribution of surgical diagnoses that characterized acute intestinal obstruction among adults that were followed-up at study sites during the study period are as shown in table 1 below:

**Table 1:** Reported surgical diagnoses that characterised acute intestinal obstruction among adults seen at Dar es Salaam public regional referral hospitals (December 2021-July 2022).

| Surgical diagnoses | Frequency* | Odds Ratio | 95% C.I of the odds ratio |
| --- | --- | --- | --- |
| Adhesions | 98 | 5.7 | 4.3 – 7.1 |
| Abdominal tumors | 20 | 1.0 | 1.0 – 1.1 |
| Hernia | 48 | 2.0 | 1.0 – 6.2 |
| Volvulus | 37 | 4.2 | 3.1 - 5.0 |

\*The sum does not add up to the total sample size as some patients presented with > 1 surgical diagnosis.

Surgical outcomes of intestinal obstruction among the study participants were also analysed. They are reported in table no. 2 the underneath:

**Table 2:** Surgical outcomes of adult patients with intestinal obstruction admitted at Dar es Salaam regional referral hospitals (Dec 2021 - July 2022).

| Observed Surgical Outcomes | Frequency (%) |
| --- | --- |
| Discharged home | 173 (76%) |
| Surgical site infection* | 20 (9%) |
| Wound dehiscence | 2 (0.9%) |
| Anastomotic leak | 1 (0.4%) |
| Referred to another hospital | 8 (3.5) |
| Dead | 12 (5.3%) |
| Unknown** | 12 (5.3%) |

**NOTE:** \* All surgical site infections observed in this study were superficial.
\*\*The figure includes those who absconded from the hospital & those who were transferred to another department within the same hospital.

To clear doubts about potential differences in surgical outcomes among the public referral facilities in Dar es Salaam, we also tested the speculation separately as shown in table no. 3 below:

**Table 3:**
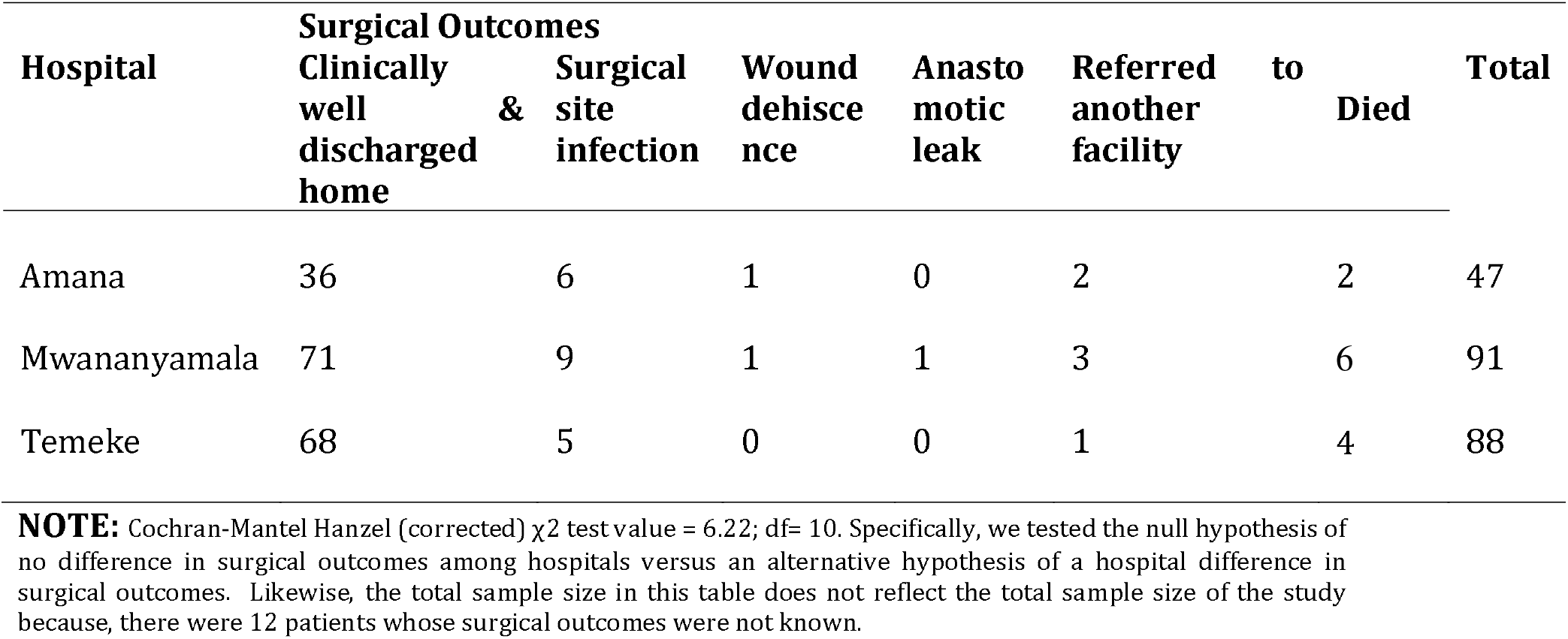
Surgical outcomes by hospital of patients with intestinal obstruction observed in Dar es Salaam regional referral hospitals (December 2021 - July 2022).

The tests for both confounders and interactions were also done but yielded non-significant findings at 5% level.

## DISCUSSION

We observed adhesions as the commonest surgical diagnosis behind intestinal obstruction during the study period. Specifically, those who had adhesions were more likely to get acute intestinal obstruction compared to those who did not all other factors in check. Our study findings are relatively unique in lieu of the fact that adhesions have been reported as the most significant antecedent for acute intestinal obstruction only from Western literature (3, 12, 13, 19, 20). However, from a strict clinical sense, our finding on adhesions may either reflect the silent change in epidemiology of insults for acute intestinal obstruction in this study population or equally likely be a result of referral bias. One should underlie the fact that our study settings were all at 3^rd^ level of referral health system in Tanzania. It is possible that most hernias (that are commonest reported antecedents for acute intestinal obstruction from the developing world) were either managed at lower level of referral facilities (e.g. district hospitals), therefore preventing the occurrence of some surgical diagnoses of acute intestinal obstruction currently studied. Should that be the case, we can safely assume the relatively lower frequency of hernia (to adhesions) observed as a cause of acute intestinal obstruction, to have been referral bias of irreducible hernias only. Our current suggestion follows new reports that also caution about the changing epidemiology of causes for acute intestinal obstruction from the developing world (21), including several from Africa before (22-24).

Likewise, abdominal tumors, hernia and volvulus were found to be statistically significant associated with intestinal obstruction. In volvulus, there is twisting of the bowels (25). Twisting of the bowels results into obstruction of the lumen; preventing the intestinal contents to move distal to the site of obstruction, and defined itself as intestinal obstruction, either partial or complete. Several studies have also shown the same pattern though differing in the estimates of the odds. Otherwise, previous studies in similar locations revealed the same findings (3, 11, 14).

We observed male preponderance (male: female =3:1) in this study population. Several studies are concordant with this finding in literature (3, 8, 9, 14, 26-29). There are several biological plausible explanations for this observation. For instance, males have increased risk towards development of inguinal hernia as well as prevalent tobacco use compared to females (1, 6, 8). For tobacco use, there appears to be an association between collagen weakness and exposure to tobacco chemicals, and therefore it may partly explain why male preponderance (1). However, detailed explanations regarding this aspect were beyond the objectives of this study. Future studies on the same agenda could also benefit science if they will also assess causal factors towards adult intestinal obstruction.

The mortality rate of more than one in twenty (5.3%) that was observed in the current study needs further scrutiny. Previous studies have yielded mixed results with respect to mortality after bowel surgeries. Mortality rates reported in literature varies widely from 0% to 42.2% (30-33). Several factors have been in place to explain the wide variation in mortality indices across studies. For instance, old age and ageing process in general has been the commonest predictive factor for mortality (31-33). It is debatable if current surgical advances in the management of intestinal obstruction benefit *senior citizens*. Management of bowel obstruction to seniors normally entails physicians/surgeons higher index of suspicion – as the condition tends to present with atypical features, that delays the diagnosis and hence likely to be in late presentation among seniors, and therefore with acute life-threatening complications at presentation, thereby potentially worse outcomes. However, the concept here should not be to defer surgical approaches whenever necessary to seniors, but rather to motivate more research about surgical conditions to this specific population group. There is clear evidence that advanced age is also associated with a number of health benefits (34, 35). There are mounting evidence in published literature that show senior citizens to constitute a significant portion not only of Africa’s population (36), but also of patients in need of care in parts of Africa’s health system (37). However, there are potential indications that clearly indicates bias against seniors’ health needs and demands, both from surgical and medical domains. (38-42) It is thus, a special call of interest to surgeons, healthcare workers and clinician-researchers in general; to consider the mortality risks as a *burden of proof* for possible link with the rising ageing population in Africa.

The fact that about one in twenty of the studied patients had unknown surgical outcomes needs further studies. This is because, there are likely multiple explanations (e.g. failure to settle the bills due to financial limitations) to account for that finding. It is therefore recommended that future studies in surgical outcomes of patients in the study settings to include socio-demographic as well as socio-economic variables as we speculate them to have been potential confounding factors behind unknown surgical outcomes in this study population.

Our study was designed in a prospective fashion and hence findings unlikely to be affected by recall biases compared to other similar works in the same topic. Besides, we did not have an upper age limit in our followed-up adults. Moreover, we studied all patients, and hence our estimates are unlikely to be associated with sampling errors prominent in most clinical research outputs in similar settings. In contrast, we also noted through our analyses, that odds ratios were used, instead of the more preferable absolute risks in clinical research. However, to the best of our thinking, we considered the malady (intestinal obstruction) to be *a relative rare pathology* in our settings, and hence our logical assumptions of odds ratios to approximate the risk ratios. It is common knowledge in clinical research, that in stances of rare events, odds ratios approximate risk ratios to the extent of being used interchangeably without much damages to the information and/or inferences made (43).

## Data Availability

All data produced in the present study are available upon reasonable request to the authors

## Acknowledgement

Authors wish to acknowledge material and emotional support from the staff in the departments of surgery at Amana, Mwananyamala and Temeke regional referral hospitals. Likewise, we send our heartfelt appreciation to the members of academic and administrative staff at Hubert Kairuki Memorial University in Dar es Salaam for their constructive supervision. Besides, we wish to convey our heartfelt vote of thanks to I-Katch Technology Ltd based COFFEESHOP RESEARCH & SCIENTIFIC WRITING programme. It formed the basis and motivation to design and publish this article, out of nearly infinite surgical research topics. We also wish to extend our vote of special thanks to management of Cafes Aroma and Central park in downtown Dar es Salaam. These two cafes situated within Barack Obama road facilitated a conducive environment for data entry, analysis and manuscript writings, some of which were done during their peak business hours. Lastly, we wish to eternally appreciate all our patients who participated in this study. It is through their willingness to participate voluntarily this article finds its strength.

## Conflict of interest statement

All authors declare to have no conflict of interest in the design, analysis and final reporting of this research findings

## Notes

### Competing Interest Statement

The authors have declared no competing interest.

### Funding Statement

This study did not receive any funding

### Author Declarations

Institutional Research Ethics Committee Hubert Kairuki Memorial University

